# Lactate arterial-central venous gradient among COVID-19 patients in ICU: a potential tool in the clinical practice

**DOI:** 10.1101/2020.05.08.20095042

**Authors:** Giuseppe Nardi, Gianfranco Sanson, Lucia Tassinari, Giovanna Guiotto, Antonella Potalivo, Jonathan Montomoli, Fernando Schiraldi

## Abstract

**Objective:** In physiological conditions arterial blood lactate concentration is equal or lower than central venous. The aim of this study was to explore if the COVID-19 induced lung cells damage was mirrored by an arterial lactatemia higher than the central venous one; then if the administration of immunosuppressant drugs (i.e. canakinumab) could normalize such abnormal lactate a-cv difference.

**Methods:** Prospective cohort study started on March 25^th^ 2020 for a duration of 10 days, enrolling 21 patients affected by severe COVID-19 pneumonia undergoing mechanical ventilation consecutively admitted to the ICU of the Rimini Hospital, Italy.

Arterial and central venous blood samples were contemporary collected to calculate the difference between arterial and central venous lactate (Delta a-cv lactate) concentration within 24 h from tracheal intubation (T_0_), and 24 hours after canakinumab administration (T_1_).

**Results:** At the T_0_ 19/21 (90.5%) patients showed a pathologic Delta a-cv lactate (median 0.22 mmol/L; IQR 0.07–0.36), suggesting severe inflammation. In the 13 patients undergoing canakinumab administration, at the T_1_ Delta a-cv lactate decreased in 92.3% of cases, being the decrease statistically significant (T_0_: median 0.15, IQR 0.07–0.25 mmol/L; T_1_: median −0.01, IQR 0.09–0.04 mmol/L; p=0.002).

**Conclusion:** A reversed Delta a-cv lactate is likely to be one of the effects of COVID-19 related cytokine storm, that could reflect a derangement in the lung cells mitochondrial metabolism induced by inflammation or other uncoupling mediators. Delta a-cv lactate decrease may reflect the anti-inflammatory activity of canakinumab. Our preliminary findings need to be confirmed by larger outcome studies.

## MAIN TEXT

## Background

The role of serial lactate determinations in critically ill patients is well agreed and may be useful in tailoring the therapy in many different diseases [1-3]. There is consensus about the two mainly observed forms of raised blood lactate concentration: lactic acidosis due to O_2_-demand/DO_2_ mismatch and hyperlactatemia with near-normal arterial pH, the latter substantially linked to hypermetabolic stress, or inherited desease [4].

In physiological conditions arterial blood lactate concentration is equal or lower than central venous (Delta a-cv lactate, normal value ≤ 0 mmol/L) [5]. Less is known about the meaning of a Delta a-cv lactate reversal (a-cv difference > 0 mmol/L), which could reflect a derangement in the lung cells mitochondrial metabolism induced by inflammation or other uncoupling mediators, likely to be responsible for the large lung parenchymal disruption (acute respiratory distress syndrome [ARDS] - like) [6]. Indeed, patients affected by severe Coronavirus disease 2019 (COVID-19) pneumonia usually present with a first phase of disease characterized by acute viremia (fever, cough, myalgia), followed by a second one, where lung injury is mainly caused by severe inflammation due to a massive release of cytokines (‘cytokines storm’) [7]. Administration of anti-inflammatory drugs (immunosuppressive monoclonal antibodies) such as tocilizumab and canakinumab have been proposed to limit the ‘storm effect’ and therefore reduce lung damage [8]. Canakinumab is a monoclonal antibody targeted at interleukin-1 (IL-1) beta which is used as treatment of the juvenile idiopathic arthritis and other autoinflammatory conditions [9-12]. Canakinumab is a relatively new biologic agent and so far its clinical use has been limited to autoimmune diseases. Our hypothesis is that the lung injury could be well depicted by an abnormal lactate a-cv gradient, while the anti-inflammatory effect of canakinumab, if any, could be mirrored by a reduction of that parameter. To our knowledge, no previous studies have examined the correlation between the lactate a-cv gradient and the use of Canakinumab.

Therefore, we performed the present study with the following aims: to describe the Delta a-cv lactate in a population of patients affected by severe COVID-19 pneumonia (early warning tool); to evaluate if the reduction/normalization of the Delta a-cv lactate was positively affected by canakinumab administration (early response tool).

## Methods

### Study design, setting and population

This is a preliminary report of the CANASCOV study (CANAkinumab Study on COronaVirus pneumonia). The study was conducted in the intensive care unit (ICU) of the Infermi Hospital, Rimini, Italy started on March 25^th^ 2020 and ended on April 3^rd^, 2020 for a total duration of 10 days.

All adult (age ≥ 18 years) patients consecutively admitted to the ICU with severe COVID-19 pneumonia who required sedation and invasive mechanical ventilation were considered for the inclusion. In the event that the baseline coupled arterial-venous lactate sampling was not obtained within 24 h from endotracheal intubation the patient was excluded. Suspected or confirmed bacterial infection (ventilator-associated pneumonia; urinary tract infection; positive blood culture; white blood cells > 12.000; procalcitonin > 1), renal failure (estimated glomerular filtrate rate < 60 mL/min/1.73m^2^) [13] or impaired liver function (liver enzyme increase more than five times the normal values) were considered contraindication to administration of canakinumab and therefore patients with any of the previous conditions contributed only with the baseline measurement of the Delta a-cv lactate.

### Data collection

At ICU admission, general patient demographic information was collected and were computed the Simplified Acute Physiology Score (SAPS II) [14] and the Brixia-score [15]. The Brixia-score is a method developed to assess the extent of lung damage in Covid patients from the chest radiogram. Briefly, the lungs are divided into three sectors on the transverse plan; to each of the obtained six sectors a score is assigned, ranging from 0 (no alteration) to 3 (interstitial-alveolar infiltrates). The score assigned to each sector is summed up and the overall score obtained may range from 0 to 18. All patients were treated with hydroxychloroquine (Plaquenil, 200 mg twice a day). Steroids were prescribed only after 7 days from hospital admission to avoid administration of immunosuppressive drugs during the acute viremic phase. Administration of steroids and antiviral drugs were also documented.

Within 24 h after tracheal intubation, one arterial and one central venous blood sample were contemporary collected by two operators through an arterial catheter and a central venous catheter with the distal tip placed in the lower third of the superior vena cava, respectively. Both samples were analyzed with an Automatic QC gas Analyzer (Siemens, Munich, Germany) and, the difference between arterial and central venous lactate (Delta a-cv lactate) concentration were calculated. PaO_2_ and PaCO_2_ were also measured and PaO_2_/FiO_2_ ratio was determined. Serum interleukin 6 (IL-6, pg/ml), creatinine (mg/dl) and alanine aminotransferase (ALT, U/L) levels were also measured at the same time. Since the laboratory simply reported IL-6 as ‘> 1000’ when its concentration exceeded this threshold, a concentration of 1000 pg/ml was considered for analysis purposes in these cases. Canakinumab (Ilaris – Novartis) 300 mg was administered by subcutaneous route in patients submitted to mechanical ventilation because of severe pneumonia caused by the COVID-19. Delta a-cv lactate was reassessed 24 h after drug administration in patients treated with canakinumab.

### Ethical considerations

Daily arterial and central venous blood samplings are part of the standard clinical practice for patient’s hemodynamic and oxyphoretic assessment. Therefore, no blood sampling was performed specifically for the study purpose. The current study was conducted according to the ethical principles of the Declaration of Helsinki. The study was approved by the Hospital Ethical Board. The research did not affect any clinical decision making in a patient’s care. The registration number of the present study was NCT04348448.

### Data analysis

The nominal variables were described as numbers and percentages, the continuous variables as medians and interquartile ranges (IQRs). Unadjusted comparisons between groups were investigated through Fisher’s exact test, Mann-Whitney U-test for independent samples, or Wilcoxon rank for matched samples, as appropriate. Bivariate association between Delta a-cv lactate and the other study variables was investigated with Pearson’s correlation coefficient (*r*). Positive or negative correlation strengths were interpreted as follows: 0–0.30: negligible; 0.30–0.50: low; 0.50–0.70: moderate; 0.70–0.90: high; 0.90–1: very high [16]. For all tests, statistical significance was set at an alpha level of p=0.05. All statistical analyses were performed using the software IBM SPSS Statistics, version 24.0 (Armonk, NY, US: IBM Corp.).

## Results

During the study period, 23 patients suffering from COVID-related pneumonia were admitted to the ICU. All patients but two were submitted to a synchronous coupled arterial and central venous sampling for blood gas analysis. Overall, the study population was constituted by 21 patients (18 males, 85.7%), whose main baseline characteristics are described in Table 1.

**Table 1.** Baseline characteristics on ICU admission of the enrolled population

| Variable | Median (IQR) |
| --- | --- |
| Age (years) | 66.0 (55.0–71.0) |
| Charlson comorbidity index | 2.0 (1.0–3.0) |
| SAPS II score | 37.0 (29.0–41.0) |
| Brixia score | 10.0 (9.0–11.0) |
| Interleukin 6 (pg/mL) | 40.3 (10.2–519.0) |
| Alanine aminotransferasi (U/L) | 32.0 (21.0–83.0) |
| Creatinine (mg/dL) | 0.75 (0.59–0.85) |
| PaO<sub>2</sub>/FiO<sub>2</sub> ratio | 195.0 (156.0–218.0) |
| PaCO<sub>2</sub> mmHg | 51.7 (41.6–55.6) |
| Arterial lactate (mmol/L) | 1.65 (0.97–2.05) |
| Central venous lactate (mmol/L) | 1.44 (0.76–1.83) |
| Delta a-cv lactate (mmol/L) | 0.15 (0.07–0.24) |

ICU: intensive care unit; IQR: interquartile range; SAPS II: Simplified Acute Physiology Score; a-cv: arterial-central venous

The median Delta a-cv lactate on admission was 0.22 mmol/L (IQR: 0.07–0.36) in the overall population. Among the 21 patients, 19 patients (90,5%) showed an inverted Delta a-cv lactate with a higher lactate level in the arterial blood than in central venous one ranging between 0.05 to 0.51 mmol/L. No statistically significant difference was showed in Delta a-cv lactate after stratification of patients according to patients’ sex and administrations of steroids and antiviral drugs. Delta a-cv lactate showed a statistically significant low positive correlation with AST (*r*=0.470; p=0.032), whilst no further significant correlations were found with severity scores, demographic, comorbidity, and laboratory variables.

Thirteen patients (61.9%) were treated with canakinumab. After the treatment, Delta a-cv lactate decreased in all but one patients (92.3%), with eight out of 13 patients (61.5%) presenting a normalization of Delta a-cv lactate (Table 2). Delta a-cv lactate decrease between T0 and T1 was statistically significant (T_0_: median 0.15, IQR 0.07–0.25 mmol/L; T_1_: median −0.01, IQR −0.09–0.04 mmol/L; p=0.002; Figure 1).

**Table 2.** Delta a-cv lactate variation in 13 patients treated with Canakinumab

| Sex | Age | Delta a-cv lactate |  |  |
| --- | --- | --- | --- | --- |
|  |  | T<sub>0</sub> | T<sub>1</sub> | T<sub>0</sub> – T<sub>1</sub> |
| Male | 32 | 0.26 | -0.77 | -1.03 |
| Male | 64 | 0.41 | -0.11 | -0.52 |
| Male | 54 | 0.51 | 0.02 | -0.49 |
| Male | 59 | 0.24 | -0.14 | -0.38 |
| Male | 72 | 0.24 | -0.03 | -0.27 |
| Male | 64 | 0.19 | -0.06 | -0.25 |
| Male | 67 | 0.24 | 0.05 | -0.19 |
| Male | 65 | 0.15 | -0.01 | -0.16 |
| Male | 55 | 0.36 | 0.23 | -0.13 |
| Male | 69 | 0.11 | 0.01 | -0.10 |
| Male | 54 | 0.07 | -0.03 | -0.10 |
| Male | 67 | 0.06 | 0.00 | -0.06 |
| Male | 66 | 0.05 | 0.11 | 0.06 |

T<sub>0</sub>: before canakinumab administration; T<sub>1</sub>: after canakinumab administration.

**Figure 1.**
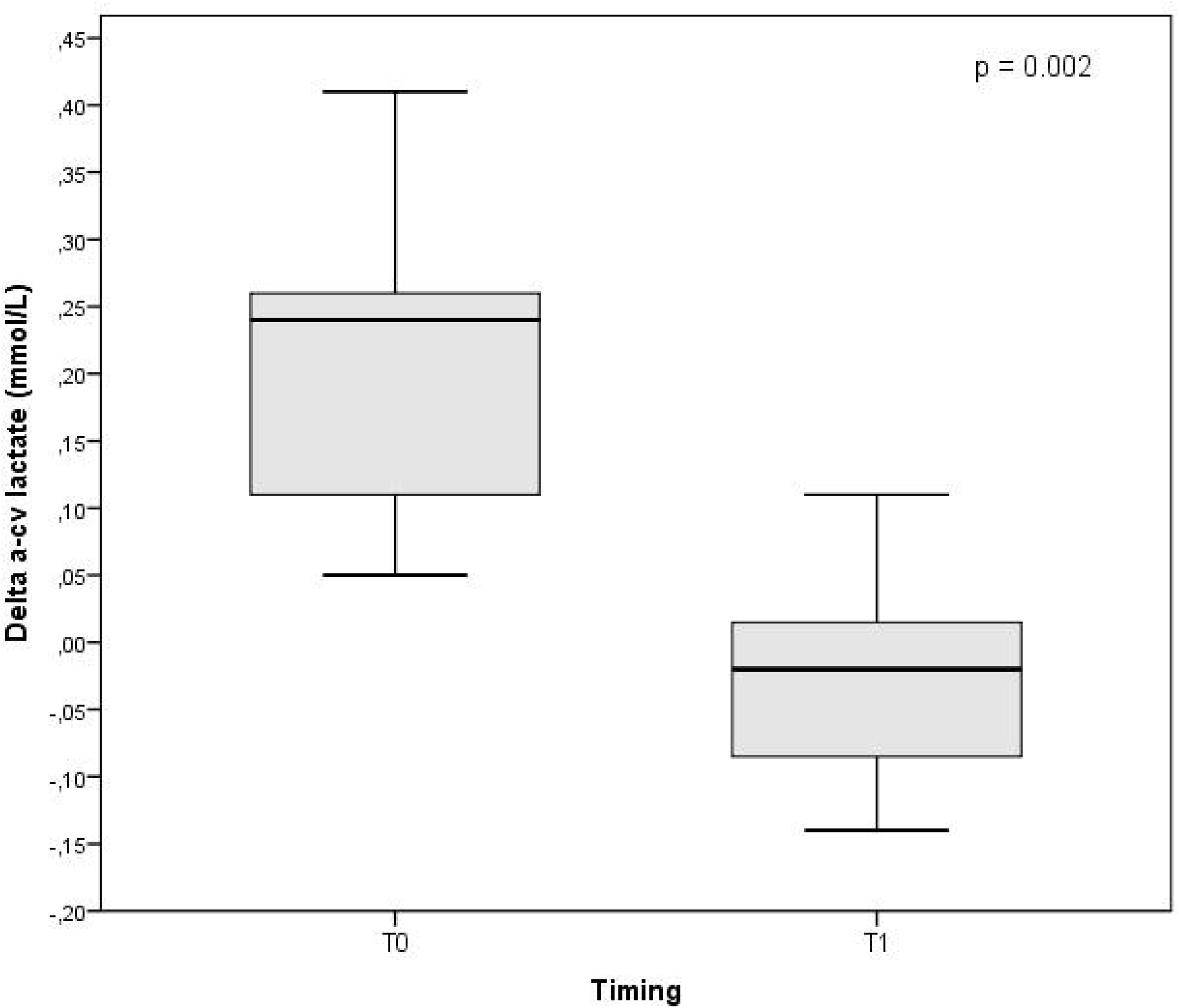

## Discussion

The study findings suggest that patients admitted to the ICU with COVID-19 pneumonia are likely to have an inverted Delta a-cv lactate that may be an early marker of the ongoing biological damage mediated by the inflammation virus-related. Moreover, the significant improvement of the Delta a-cv lactate found in the majority of patients treated with canakinumab could be in interpreted as a favorable, early effect of the administered medication, supporting the hypothesis of a reduction of cytokines-related severe inflammation at the lung level. However, the present study is a small observational study and the lack of a linear correlation between the Delta a-cv lactate and IL-6 levels requires caution with the interpretation of the study results calling for further studies aimed to provide a better understanding on the role of immunosuppressive drugs in the treatment of the COVID-19 pneumonia. In addition, the use of immunosuppressive monoclonal antibodies is associated to potential risks of liver injury and decreased patient’s immunocompetence. Therefore, more data and a larger sample of patients are needed to analyze in deep the effectiveness of these anti-inflammatory drugs.

As the COVID outbreak is becoming a real ‘stress-test’ for Healthcare Systems all over the world, many different pharmacologic strategies are under investigation against strong outcome indicators, such as in-hospital death or ICU length of stay. In this context, the availability of predictors able to identify early, quickly and without additional costs patients in whom the infection is having a greater impact is pivotal. To the best of our knowledge, this is the first study considering Delta a-cv lactate assessment in mechanically ventilated ICU patients with severe COVID-19 pneumonia. Interestingly, Delta a-cv lactate data can be collected routinely bedside and almost for free, even in an overwhelmed ICU context, as the COVID-19 patients in ICU are usually submitted to central venous and arterial cannulation to allow hemodynamic monitoring and blood gas analysis.

### Limitations

Our preliminary report has several methodologic limitations, that should be considered when interpreting our results. The study had an observational design with descriptive and correlational aims and enrolled a rather small sample in a single center. Therefore, the possible causative relationship between canakinumab administration and Delta a-cv lactate decrease, as well as the predictive power of this variables on patients’ key outcomes, should be tested in larger populations and after considering relevant confounders. Moreover, although our findings may be explained by the effect of the administered drug, a direct comparison of the trend of the Delta a-cv lactate in a group of patients receiving the standard of care was hampered by the lack of a control group.

## Conclusion

The serial evaluation of Delta a-cv lactate trends seems to be a fast reliable tool to get a further insight about the lung biological disarray, perhaps even useful to better tailor the therapy in COVID patients. Nevertheless, our data can only suggest that the use of canakinumab may normalize the Delta a-cv lactate, thus supporting the hypothesis of a reduction of inflammation at the lung level. Therefore, our results should be only considered as a contribution to a better understanding of the physiopathology of COVID-19 induced lung injury.

## Data Availability

Data could be available upon request

## Acknowledgments

The authors thank the whole staff of the study ICU for their precious collaboration in data collection.

## Author Contributions Statement

GN and FS conceived the study; GN and GS designed the study; AP, JM and FT performed data acquisition; GN and GS analyzed the data; GG, GN, JM and FS interpreted the data; GG, GN, GS and FS drafted the manuscript; all Authors revised critically the manuscript; all Authors took responsibility for the paper as a whole, ensuring that questions related to the accuracy or integrity of any part of the work were appropriately investigated and resolved; all authors have seen and approved the manuscript.

